# Socio-cultural practices, dietary and nutrition patterns, economic and vitamin-D deficiency status of pulmonary tuberculosis patients (PTB) of tribal and urban population of India: An Explanatory Model Interview Catalogue (EMIC)

**DOI:** 10.1101/2023.07.21.23292873

**Authors:** Sushanta Kumar Barik, Jyotirmayee Turuk, Meenu Singh, Sidhartha Giri, Sanghamitra Pati

## Abstract

This study emphasizes on the implication and benefits of an Explanatory Model Interview Catalogue (EMIC) on the collective information of the socio-cultural status, diet and nutrition pattern, economic status, and treatment options through proper counselling of pulmonary tuberculosis patients (PTB) during the acquiring of the tuberculosis (TB) in tribal and urban population. TB is one of leading cause of death in India. It is necessary to find out progression of PTB associated with socio-cultural practices, dietary and nutrition patterns, economic status, and Vitamin-D deficiency among TB patients of tribal and urban population for better management of tuberculosis patients.

Therefore, EMIC developed will be applied for collection of details on diet and nutrition, types of occupation, social status, types of complementary consumable additives, economic status, types of community and languages, Vitamin-D status, treatment details, quality of life in newly diagnosed PTB patients in state level, district level hospitals and medical research institutions.

## Background

### Description of the topic

India living with 8.6% of tribal population creating a bridge gap between tribal and non-tribal population in health care (Bisai 2014). The focus on the health of the tribal population is very important as they are the socioeconomically deprived sections in India. Tribal health care system is based on their traditional knowledge system and the promotion as well as execution of the health care system is very essential in every level of tribal population of India (M Kumar 2020). The prevalence of the PTB in different studies among the tribal population was estimated. The study will address the need for a standardized method for screening of resistant TB, improvement of methodological strategies, health seeking behaviour of tribal people, chest symptomatic and essential for increase the assessment of health care services (Thomas 2015). It has seen that one of the contributing factors for causation of TB progression of latent TB infection to active disease, the risk of drug toxicity, relapse, and death (Padmapriyadarsini 2016). Vitamin-D deficiency was prevalent among the PTB patients with extensive lesions and severe clinical symptoms (Jaimni 2021). One of the studies conferred that undernutrition was observed among the PTB patients and essential need of proper management and counselling of TB patients (Shukla 2019). In India, the adult tribals are particularly vulnerable to undernutrition, because they are geographically isolated, socioeconomically disadvantageous with inadequate health facilities available (Rao 2006). The prevalence of TB is associated with the multidimensional poverty of the population in India (Sarin 2017; Pathak 2021). Dietary counselling improves the quality of life of PTB patients (Pajanivel 2022). Supplementation of dietary micronutrient beneficially increase the cure rate of PTB patients (Xiong 2020). Undernutrition and TB have a bidirectional relationship among the various population of India and must be a situational analysis to way to froward by improving the nutritional status to decrease the risk of disease transmission (Padmapriyadarshini 2016). Early studies investigation was confirmed the relationship between the nutrition and immune response play an important role to fight infection and reduce the risk of disease progression. Nutritional deficiencies and excesses in the body of human beings always influence the various components of the immune system. Deficiencies of dietary proteins, amino acids, vitamins A, E, B6 and folate, trace elements like zinc, copper, selenium etc. were associated with reduced immune competence towards pathogenic infection (Chandra 1986). Dietary vitamin -D and sunlight exposure on the production of vitamin-D is very essential to fight the infection for *Mycobacterium tuberculosis*. Insufficiency in Vitamin-D in the body of human beings were highly prevalent with PTB patients (Desai 2012).

The importance of health education along with the collection of nutritional indicators, immune level, and quality of life of the PTB patients is very essential. Thus, a comprehensive study of the socio-cultural practices, dietary and nutrition patterns, economic status, and vitamin-D deficiency of PTB patients of tribal and urban population of India is very essential in a program-based manner using a suitable EMIC based systematic protocol.

### A brief knowledge of the review protocol what is already known

To focus on the structural forces on TB related stigma was very important to understand the spread of the TB in a population level. TB related stigma was primarily associated with the poverty, malnutrition, and HIV coinfection. The basic instrument in epidemiology is the Explanatory Model Interview Catalogue (EMIC) based on specific demographic and sociocultural settings for collection of information on TB related stigma. The structural epidemiology influences shape of the TB related stigma. The explanation based on interview model could drive out the unhealthy related TB stigma to the community. The recruitments of study participants were always welcomed by the interviewers, and spreading the knowledge of TB in a community (Coreil 2010). Systematic mapping of epidemiological structure by various methods was completely known in TB related stigma. Various conceptual frame work was designed to understand the TB stigma. The collection of social and structural determinants of health for measurement of burden of TB could be a special focus in a population level. The systematic mapping of the epidemiological factors, strengthen the scenario of TB (Craig 2017). TB was associated with psycho-socioeconomic quality of life and could be recovered by successful antitubercular treatment. The TB related quality of life and its impact on treatment could be explained through EMIC (Datta 2020). The delayed recognition of TB impacts the social and cultural life of TB patients. The cultural epidemiological approach through EMIC based was useful for explaining the treatment adherence, cause of default in various epidemiological determinants and its impact on gender specific behaviour on TB (Gosoniu 2008). In a TB clinic, the semi structured EMIC was administered to study the cultural epidemiology of TB patients. Loss to diagnostic follow-up and delay in health seeking behaviour of TB patients were observed (Mhalu 2019). The development of innovative TB reduction strategy is essential in rural and urban areas. Thus, an appropriate TB stigma related information would require using the EMIC to disseminate the health status of the people (Oladele 2020). Social stigma is an important barrier of TB transmission associated with low level education and socio-economic status. Measurement of social stigma among TB patients through EMIC model could reduce the TB burden and transmission in community (Shah 2020). Socio-cultural determinants of TB related stigma are associated with the life style of men and women and could evaluated by EMIC (Somma 2008).

As TB control is a global challenge, it is better understood to take epidemiological context of TB burden and transmission in a population level. The EMIC will be helpful in reduction of TB in the institutional, hospital and community level. The use of EMIC will improve the patients right to confidentiality of care in a hospital, attention of the patient’s report to media, social isolation, negative stereotyping about TB, immigration regulations, government programmes, public health services etc. We need to characterize the structure of the socio-cultural practices, dietary and nutrition patterns, economic status, and vitamin-D deficiency of PTB patients of tribal and urban population of India through an Explanatory Model Interview Catalogue (EMIC) to reduces the burden of TB stigma.

### Why the study design and targets are essential?

The different tribal groups are in a different phase of socioeconomic development in many states of India. It would be very essential to generate the tribe specific data, giving most priority to tribal groups based on development and vulnerability and start-up of several development programs in several districts of India (Saha 2016). Transforming health system in the tribal areas will be great achievement for equality in India. The innovation and expansion of provision of health services, development of human resources for health services, management of cash transfer system, closing the gap between the maternal and child nutrition support of schedule tribes will improve the health system in India (Thomas 2015). Many tribals in India are live in adverse environmental conditions and are exposed to several types of infectious diseases throughout the year. The improvement of the health care, diet and nutrition, economic facilities will be pathbreaking in the tribal population, which are in verge of being extinct (Meher 2007). The TB stigma is also cause of spread of TB among close contacts is due to fear of social isolation, fear of losing social status, verbal abuse, gossip, neglect from family and failed marriage prospect (Thomas 2021). Thus, a model-based investigation through the EMIC based protocol would assist the Revised National Tuberculosis Control Program (RNTCP) in India. As TB is one of the oldest communicable diseases digging deeply in the soul of the mankind of the world, it is very important to diminish the traditional methods and improve modern health care system for better assessment of tribal in different states of India (Negi 2021). Thus, a systematic protocol on the socio-cultural status, diet, nutrition pattern, economic status, and treatment option is designed to assess the sociodemographic and sociocultural aspects of tribal population.

### Why the study of social determinants is so important?

The social determinants are the key risk factor of TB which is evident from the published material (Hargreaves 2011). As per the evidences of TB control program in low- and middle-income countries not only the diagnosis and treatment options are required for TB management but also the study of social determinants of TB. The practical ides on in strengthening of tuberculosis control program needs to be developed according to the World Health Organization Commission on the current understanding of the social determinants of TB. Social protection and urban planning would have strengthened the tuberculosis control program. The explosion of research on social health, human rights, and poverty reduction strategy on socioeconomic inequalities of opportunity in the individual’s health is essential to study the social determinants of TB. The emphasis on urban regeneration, housing design, slum upgradation is the key work on the social determinants of TB. The proper dietary practice and nutritional status also plays a significant role during treatment of TB (Gurung 2018). Thus, the proper counselling of the assessment of dietary intake and nutritional status should be done periodically to reduce the burden of social determinants of TB patients. The nutritional support should be given to severely malnourished to drive out the social and economic issues of TB patients. The aggressive role of health care workers also focuses the key social determinants of TB patients to end TB in India by 2025.

### Why the study of cultural determinants is so important?

Socio-cultural changes are an important issue in the developmental changes and pattern of life of the tribal and urban population of India. Tribals of India are restricted in the social, political, and legal relationship in a family. Tribal population of India are much varied and diversified. The cultural patterns are varying from one tribe to another tribe in India. The tribals are rich in cultural diversity, traditional customs, festivals, and vibrant life with dances. In general population, cultural related of TB is associated with illness experience, behaviour associated with culture, TB transmission in a gender specific manner. Women were generally less aware of the cause of TB than men due to a cultural metaphor for fate called *Karma*. Gender specific acquire of TB is the common cultural practice of human beings in a community. The documentation and quantification relevant to cultural aspects is very essential for policy makers and program implementers in the sensitive area of TB. EMIC based structural interviews were conducted on the specified health problems of the human beings in a community (Atre 2004). Global cultural variations are concerned in response to TB stigma. The study of cultural variation should be addressed for reducing TB stigma and treatment adherence of improvement in the human population (Chang 2014). The consumption of alcohol considered as ‘‘a holy drink’’ and a strong as well as traditional cultural beliefs of Indian tribes (Chaturvedi 2019). The study of cultural factors is most important compared to the structural factors for the TB patients in a hospitality or community. It is necessary to address the cultural determinants to perform the research and draw an inference of the study (Chaturvedi 2011). Thus, a more specific study on the cultural determinants for TB patients of various tribes of India through EMIC is very essential.

### Why the study of dietary and nutrition pattern determinants is so important?

Undernutrition is a leading risk factor of TB incidence and is a common risk factor for mortality of TB patients in India. Therefore, the nutritional supplementation on TB incidence is very important through the reducing activation of TB by improvement of nutritional status (RATIONS) study in a population level (Bhargava 2021). Malnutrition is the secondary immunodeficiency of the human being’s susceptibility to Mtb. Various factors like nutrient malabsorption, micronutrient, and macronutrient malabsorption, altered metabolism, protein energy malnutrition are the important issues of the human beings for a risk of infection to TB. Supplementation of various nutrients is a novel approach for fast recovery of TB patients (Gupta 2009). Dietary and nutrition practice through counselling is an effective assessment for TB patients. The improvement of nutrition status is very essential from the time of registration to the time of study in case of severely malnourished TB patients (Gurung 2018). Therefore, there is an urgent need of proper dietary and nutrition counselling of severely nutrition deficiency TB patients admitted in hospitals or living at home will be required for management of life style. Thus, the implementation of EMIC model for TB patients for assessment of the diets and nutrients could improve the quality of life.

### Why the study of economic status determinants is so important?

The analysis of magnitude and pattern of TB related cost is very important for each people suffering from TB. The economic status of the TB patients persists until the completion of the treatment. The issues of the socioeconomic conditions, income, out of pocket expenses, drug pick-up, hospitalization, additional food, medical pick up, treatment outcome, identification of post treatment symptoms etc. are associated with the living cost of the TB patients (Chatterjee 2023). The burden of TB is associated with the various socioeconomic classes of the people. The income of the patient is associated with the therapeutic episode in TB patient (Imam et al, 2021). The economic and social status of the people affects the TB patients in high burden countries to downhill their sustainable development goals (Silva, 2021). The direct and indirect cost of living of the people with TB was analysed. It is realized to India’s elimination of TB by decreasing the diagnostic delays, reducing the travel cost, improving awareness, ensuring sufficient reimbursement and search of active case finding through proper diagnosis. How much do Indians are paying for TB treatment cost was analysed (Sinha 2020). The understanding of financial burden of each TB patients and rising the health care expenditure of TB patients is an effective challenge to achieve TB free India by 2025 (Yadav 2021). Thus, the necessary steps would have taken through an EMIC model of investigation for economic stabilization to reduce the burden of TB in a programmatic manner.

### Why the study of vitamin-D deficiency status is so important?

Vitamin-D acts like a signalling molecule to modulate the activities of macrophages/monocytes, dendritic cells, T and B cells, body epithelial cells to induce inflammatory responses to infection and immune functions. Vitamin-D deficiency leads to progression of TB and could use as a potential adjunctive therapy for TB (Cai 2022). Vitamin-D association and TB disease progression was identified. Low Vitamin D levels in TB patients increased the risk of progression of the disease (Talat 2010). The impact of vitamin-D levels on the risk of TB was assessed. The risk of TB progression was studied by supplementation of Vitamin-D in dose dependent manner (Aibana, 2019). Thus, the assessment of vitamin-D is very important for the newly diagnosed TB patients until the completion of diagnosis. The most practicable way to use of the EMIC model for quick assessment of the vitamin-D status of TB patients enrolled in the hospital or living at home.

### Why the health system improvement is essential for TB patients using the protocol?

Mahatma Gandhi, father of nation, said that India lives in its villages. TB treatment and cure only be possible if door step services are provided to TB patients (Natrajan 2019). The Govt. of India has launched a scheme ‘’ Nikshaya Poshan Yojana’’(NPY) for a monthly financial incentive to improve the nutritional status of the TB patients (Kumar 2020). The health and health care delivery are an important vision of prime ministers of India. Empowering the tribal population on socio cultural practices, health care facilities, socio behavioural practices and creation of self-reliance vision of unprivileged tribal group is a step by the govt. of India on the tribal and urban areas of India (Saxena 2022). On September 9, 2022, the President of India, virtually launched the Pradhan Mantri TB Mukt Bharat Abhiyaan. To keep the better detection and treatment for TB patients, the govt. has started the doorsteps diagnosis by health care workers through TB van and sensitizing with EMIC. This suitable EMIC could help for a better information on TB diagnosis in urban and tribal population. The Government of India is planning to focus the improvement of health system for TB detection and treatment plan to end TB by 2025 in urban and tribal population level. Now, India has raised the social service and adaptation system, treatment, and nutrition value for urban, rural, and tribal TB patients. India targets for TB Mukta Bharat by 2025. Urgent actions are needed to see the effect of TB on Individuals, family and society, TB awareness campaign, failure of govt. plans, focus on health care infrastructure by improving the quality and quantity of medical facilities and doctors. A lot of variation was found in the private health sector on tuberculosis care in urban areas of India (Kwan 2018). A very strong supportive EMIC is essential to strengthen the management of TB in the urban India’s private sector too. Lancet on TB addressed to improve the quality to building a TB free world. Thus, we need a right commitment of leadership and utilization of resources to realize the quality of care for TB patients in both urban and tribal population of India.

### How EMIC will assist in the interventional study of the PTB patients?

Nutritional supplements and regular counselling could improve the weight with a greater rate of treatment success of TB patients in India (Singh 2021). But along with the additional food distribution system, home counselling, personal discussion on the treatment success of the PTB patients could be possible by EMIC is the additional benefit for the intervention study group of tribal and urban population level. In last few years, the govt. of India encouraging the management capacity of the TB program, treatment delivery and drug supply, pushing political commitment in the private sector, technical innovation for health sector improvement through EMIC for TB patients of the urban and rural sector in India. Personalized intervention will be required to improve the medication adherence in TB patients (Pradipta 2020). The importance of the EMIC could assist the policy and practice during the interventions of the PTB diagnosis and research in several private and govt. organizations at urban and tribal areas of India. The use of the EMIC could reduce the mortality and transmission of TB during interventions. Interventional study can reduce the morbidity or mortality rates, reducing the risk of ill health by identifying the key elements of the population health of tribal population of India (Mohindra 2010). The review of the standard regimens during treatment of PTB is very essential and can be included as an interventional study for each PTB patients (Grace 2019).

A strong EMIC will address the essential needs and fill up the knowledge gap in the tribal and urban population of India.

### What are the benefits and impacts of the EMIC for PTB patients of tribal and urban population level?

- EMIC will be useful in the collection of verbal information from the PTB patients through knowledge enhancement and recalling ability in the tribal or urban population.
- EMIC will have an impact on the diet and nutrition as well as life style changes in the PTB patients to make a better visibility of life in a tribal or urban community.
- EMIC will be helpful on the ethics related study on human beings acquiring any type of TB in tribal or urban population.
- EMIC utilizes the qualitative data of the study subjects of the human beings in a PTB infection in a tribal or urban population level.
- EMIC should design in a suitable manner with a greater impact on the educational knowledge, practice, and attitude of the PTB patients towards their disease management in a tribal or urban population level.
- EMIC will generally strengthen the general practices and promotion of the health literacy of the PTB patients in urban and tribal population level.
- EMIC will help the health care providers for their general practice for PTB patients in tribal and urban population level.
- EMIC will engage with health classification frame work like health promotion, disease prevention, health care and management, health protection of PTB patients in tribal and urban population level.
- EMIC will give a good readability and subject content on health frame work of PTB patients in tribal and urban population level.
- EMIC assists in the literacy skills for life survey of PTB patients in tribal and urban population level.
- EMIC will make easier for the patients and pharmacists to collect the information about their treatment in PTB patients in tribal and urban population level.
- EMIC will relieve the difficulties of obtaining reliable and enough information of PTB patients in tribal and urban population level.
- EMIC improves the readability scores of the PTB patients in English, Hindi, and other regional languages in tribal and urban population level.
- EMIC will exact support the distribution of health literacy skills in an independent manner of PTB patients in tribal and urban population level.
- EMIC improves the general literacy skills in a participation manner in every day life of current economy of PTB patients in tribal and urban population level.
- EMIC create a set of minds to accomplish everyday health tasks of PTB patients in tribal and urban population level.
- EMIC enhances the literacy proficiency by evacuating the mental disability in health tasks platform of PTB patients of tribal and urban population level.
- EMIC enhances the strong willingness of the PTB patients for preparing the future chronic disease attack in the tribal or urban population level.
- EMIC increase the knowledge of health communicators to strongly support the knowledge of PTB patients in tribal and urban population level.
- EMIC will enhance the recalling ability of the PTB patients in the tribal and urban population level.
- EMIC will enhance the verbal communication between a doctor and a PTB patient to secure daily health tasks of tribal and urban population level.
- EMIC will be useful in the acute condition cases where the PTB patients suffer from the lack of information on sociocultural status, diet and nutrition pattern, economic status in the tribal or urban population.
- EMIC will fulfil the essential needs of getting information of the PTB patients in the tribal and urban population level.
- EMIC will assist on straight of mind of the PTB patients on medical terminology acceptance in the tribal and urban population level.
- A tailor made EMIC will help the PTB patients to control their own sociocultural status, diet and nutrition pattern, economic status, and healthcare improvements in the tribal and urban population level.
- EMIC will work as a strong public health support and guidance in sociocultural status, diet and nutrition pattern, economic status, healthcare, and treatment option for PTB patients in the tribal and urban population level.
- EMIC will strongly support the better diagnosis by collecting a lot of information from the PTB patients and prevent from the delayed diagnosis.
- EMIC will be helpful in the TB stigma reduction strategies in the institutional, governmental, and structural levels in tribal and urban area.

## Methods: Criteria of the study using the EMIC for PTB patients

### Types of studies at urban and tribal population level

Criteria for considering the review protocol: We will include the EMIC protocol in the PTB patients of urban and tribal population for a qualitative study design and data collection methodology. We will collect information by conducting the structured and unstructured interviews of tribal and urban people, call up more participants and collect information of tribal and urban people by oral histories. We will analyse the qualitative and survey data of the socio-cultural practices, dietary and nutrition patterns, economic status, and vitamin D deficiency of PTB patients of tribal and urban population of India.

### Topic of Interest

Any study related to PTB in active case findings that includes the targeted screening of high-risk populations deprived in vitamin D deficiency and nutrition supplements in tribal and urban population level of India. We will focus the social and cultural characteristics of the whole tribal and urban communities in the chosen study area. We will think for a better screening of health associated with PTB of urban population and rural tribes and more focus on the improvement of the health care systems in urban and tribal population.

### Participants and populations of the PTB patients

We will include the active population of house hold contacts, whole communities, minor and other workers at their work place in the urban and tribal area of India in the age of 18 to 60 years of active PTB cases of tribal and urban population.

### Criteria of the participants

Patients with newly diagnosed pulmonary tuberculosis who fulfils the inclusion and exclusion criteria

### Inclusion criteria

1. Age 18 years to 60 years inclusive.
2. Male and Female.
3. Not pregnant or breast feeding by history.
4. if selected, subject must be willing to give venous blood 2 times and sputum samples 3/4 times for examination in study period.
5. If selected, agree to have blood stored for future studies.
6. Ability to understand and give informed consent.
7. Healthy controls of same age and sex group (people not suffering from tuberculosis or other chronic diseases like cancer and with no history of diabetes, chronic kidney disease or HIV).

#### Exclusion criteria

1. Age below 18 years or above 60 years.
2. Pregnant or breast feeding by history.
3. History of diabetes, chronic kidney disease or HIV.
4. MDR/ XDR diagnosed patient including Category II (defaulters, relapse, or re-treatment).
5. Healthy people with exposure to tuberculosis (like family members of patients) or suffering from other chronic diseases like cancer, HIV, diabetes etc.

### Study design and settings

The study will be set up in the community health care centres, primary health care centres and district hospitals of tribal and urban district of Odisha, India. All male and female active cases of PTB will be included. A questionnaire will be assessed on the social, cultural, and nutritional status of the tribal and urban population of the specified district.

### Co-morbidities of the participants

Participants in this qualitative study have a problem of upper respiratory infections or seeking illness.

### Interventions of the PTB patients

Active PTB cases of the urban and tribal communities are mandatory for improving the health facilities through a better screening by using the EMIC. The health promotion activities of PTB patients of the urban and tribal population level could be a better judgement through a proper intervention.

### Search of Methods for Identification of the study in tribal and urban areas

We made a search of the sociodemographic paper, cultural practices papers, diet and nutrition papers, vitamin D deficiency papers of previous study tribal and urban population on TB or PTB. We searched the following databases: Google scholar, MEDLINE(PubMed), Science direct, social science index, Cochrane library search (https://www.cochranelibrary.com/), Science Open Research (www.scienceopen.com) for extracting the study papers on PTB patients.

### Language translation

We will translate the English format EMIC in to the Odia or respective tribal language for easily assessment of the health care those are suffering in active PTB cases.

### Data management, analysis, and synthesis

Individual data on sociodemographic, cultural, diet and nutrition pattern of the active tuberculosis cases of the tribal and urban population level will be conducted by the EMIC. Individual data will be managed in a simple EMIC in paper based hard copy or electronic format. The data will be synthesized and analysed by the help of the statisticians and the descriptive statistics will be used for further analysis of the data and will be communicated to the physicians and health care workers of the urban and tribal areas of India.

### Control measures PTB patients

Level of awareness, advocacy, communication strategy, increasing the health education activities could assist the TB control and elimination of TB in the national program (Chinnakali 2013). Increasing the level of awareness on ‘’free treatment and adoption of the new regimens for treatment for PTB infected and drug resistant patients in urban poor slum will be a good initiative in controlling PTB. There is need to further strengthen the four strategic pillars of ‘Detect-Treat-Prevent-Build’ (DTPB) to control PTB and continues in the program laid by govt. of India in the national tuberculosis elimination program (NTEP). Now, the time must come to give more sustainable growth of tribal and urban population by expanding and focusing the TB control program likely end TB by 2025, by increasing country’s level of economic growth, building the diet and nutritional pattern, improving the primary and private health care system and gives special attention to prevent HIV and multidrug resistant TB through a designed EMIC.

## Conclusion

### EMIC implication in practice

The new strategy for EMIC should ideally be rapid, inexpensive, understandable, and verbally acceptable for PTB patients of tribal and urban population level. The intensive queries on the different aspects of the EMIC enhances the collection of the real facts of socioeconomic, cultural, nutrition and dietary status in the tribal and urban population. The extensive collection data through EMIC must applicable by the social workers and might be easily stored in a database format for the future practice in the epidemiological area of research on PTB patients of the tribal and urban population. At present, the national TB programs are well associated with the pharmaceutical and nutritional supplements to the TB patients to improve the quality of life as well as treatment outcomes, so the implementation of the EMIC helps to qualify more eligibility of the PTB patients in tribal or urban population level. The implementation of the EMIC possibly enhances the nutrition management and awareness practices in the TB research to the national media. The EMIC keep the continuity in health practices periodically accessing the nutritional status to severe the malnourished PTB patients. The implementation of the EMIC probably work like an educational support on dietary guidance to support the immune system by improving the quality of life of the PTB patients in the tribal and urban population. The EMIC implementation will update the nutritional assessment in a simplification manner throughout the treatment of PTB patients in the tribal and urban population. The implementation of the EMIC impacts upon health education of the tribal and urban population of India. The implementation of the EMIC impacts on the personalized dietary counselling on the nutritional status during the treatment of PTB through the medical practice.

### EMIC implication in research

The EMIC will assist in the research clinical settings for PTB patients in the tribal and urban population. The EMIC vigorously collect the basic needs of the tribal and urban population affected in PTB and prepare the guide line of data collection for research and publication. EMIC will assist the organization of therapeutic nutrition-based TB research through the assessment of the nutritional status of the patients and identification of the imbalance macronutrients in the diet. The EMIC could assist in the pilot project-based research on sociodemographic, economic, diet and nutrition pattern of TB/ PTB patients in various medical universities and research institutions in India. The implementation of the EMIC will works like a multitask educational health guideline in the health management research in the area of nutritional monitoring, dietary requirements, immune function, general information on quality of life, treatment complications etc.

### EMIC implication in social progress index (SPI)

The improvement of the social progress index (SPI) is very essential in the tribal and urban population level as India holds its position 110^th^ in the world. The EMIC will help the social progress of the tribal and urban population. It must assist to improve the social index score of each tribal and urban population states of India. Gradually, the lower social progress score index of a state will be converted into higher social index score progress state by utilization of the EMIC in the tribal and urban population. The EMIC will basically focus the three dimensions of human needs such as (**A) Basic needs of human beings** (i) Diet and nutrition (ii) Basic medical care (iii) Pure water (iv) sanitation (iv) Shelter and safety **(B) Wellbeing features:** (i) Access to basic knowledge (ii) Access to information (iii) Access to communication (iv) Access to health (v) Access to environmental quality **(C) Opportunity** (i) Personal rights (ii) Personal freedom (iii) Personal choice (iv) Focus on advanced education. By use of the protocol may influence the SPI of the PTB patients of the tribal and urban population states of India and assist to improve the cooperative knowledge of various primitive tribes to come out a future of living style. The EMIC must help to secure their traditional progress within a tribal community to eyes on the index value of the SPI of the country.

## Data Availability

Online

## Acknowledgement

Dr Sushanta Kumar Barik, ICMR-RA fellowship (Award letter No. 3/1/2/299/2021-Nut) on the project entitled ‘’A comparative study of the vitamin-D deficiency, vitamin-D polymorphisms in pulmonary tuberculosis patients in tribes and urban population of Odisha, India’’) by ICMR, Govt. of India is acknowledged.

## Authors Declaration

All authors declare no conflict of interest.

## Ethics statement

The study entitled **‘’**A comparative study of the vitamin-D deficiency, vitamin-D receptor polymorphisms in pulmonary tuberculosis patients in tribes and urban population of Odisha, India’’(Ref:..ICMR-RMRC/IHEC-2022/118, Date:25/7/2022) was approved by the Institutional Human Ethics Committee (IRB No:(ECR/911/Inst/OR/2017) ICMR-Regional Medical Research Centre, Bhubaneswar, Odisha, India.

## APPENDICES

### Appendix 1. Search strategies

#### PubMed (MEDLINE)

| Search | Query |
| --- | --- |
| #1 | Pulmonary Tuberculosis |
| #2 | Sociocultural features of tribal population of India |
| #3 | Dietary practices of Tribals of India |
| #4 | Vitamin D deficiency in tribal population of India |

#### Google scholar

| Search | Query |
| --- | --- |
| #1 | Pulmonary Tuberculosis |
| #2 | Sociocultural and economic burden of tribal population of India |
| #3 | Dietary practices of Tribals of India |
| #4 | Vitamin D deficiency in tribal population of India |

#### Social Science citation Index

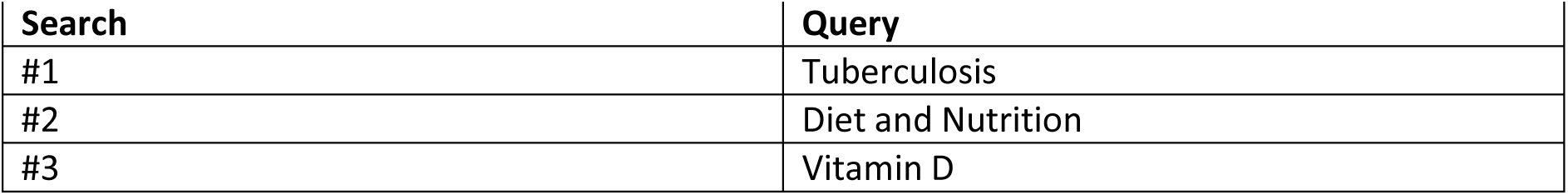

| **Search** | **Query** |
| --- | --- |
| #1 | Tuberculosis |
| #2 | Diet and Nutrition |
| #3 | Vitamin D |

#### Cochrane Library search

Published protocols and reviews on Pulmonary tuberculosis

#### Science Open Research (www.scienceopen.com)

Search articles using the words like Tuberculosis, pulmonary tuberculosis, vitamin-D in Tuberculosis patients

## DECLARATIONS OF INTEREST

All authors declare no conflicts of interest

Sources of Support:

Indian Council of Medical Research, Govt. of India

File No. 3/1/2/299/2021-Nut.

## Supplementary file: Explanatory Model Interview Catalogue (EMIC)

A. **Type of patients:** Pulmonary Tuberculosis patients
B. **Types of Population:** Tribal/ Urban
C. **Vitamin-D Uptake**: Yes/ No
D. **Vitamin-D concentration**: ……….ng/mL **E**. **After ATT completion** ng/mL

### Socio-Cultural Catalogue-1

1. **Registration Number:**
2. **Date of start of ATT:**
3. **Date of Registration :…../……/……**Treatment status at registration: On ATT ( ) Not on ATT ( )
4. **Name of study center**: ………………………………………………
5. **ATT center code**……….
6. **City :…………………State:……………….**
7. **Name of patient**:………………………**Gender:** Male( ) Female ( )
8. **Age (Years):**
9. **Date of birth:……**………
10. **Category:** General ( ) SC ( )ST ( ) OBC ( ) Others ( )
11. **Patient’s phone number**…………………………………………………
12. **Address:………………………………………………………………………………………………………………**
13. **City/Village:…….Tehsil/Taluk………………District:…………. State: ……………………..**
14. **Care givers name:…………………………………………**
15. **Care givers address and phone number: ……………………………………………………**
16. **Types of Diet**: Vegetarian or Non-vegetarian
17. **Occupation:** 1. Unemployed ( ) 2.Manual labor ( ) 3.Skilled labor( ) 4.Clerical( ) 5.Business ( ) 6.Professional ( )7. Defense and Security personal ( ) 8.Community social worker ( ) 9.Housewife ( ) 10.Student( )11.Religious/Spiritual leader ( )12.Salesman ( )13.Farmer ( ) 14.Entertainer/Musician/Artists ( ) 15.Cook/Waiter ( )16. Hotel Boy ( )17.Auto Driver ( ) 18.Longdistancetruck and busdriver ( ) 19.Vendor( )20.Technician( )21.Brothelkeeper( ) 22.Police( ) 23.Watchman( )24.Selfemployed( )25.Retiredofficial( )26.Beggar/Ragpicker( ) 27.Supervisor/Maneger( )28.Delivery Man( )29.Athlete( )30.Govt.servant( )31.Others( ) 32.Refused( ).33. Agricultural labor ( )34.non agricultural labor ( )35. Domestic servant ( ) 36.Skilled worker ( ) 37.Semi skilled worker ( )38.Petty business/large business/small shop/self-employed ( ) 39.Service(govt/pvt.)( ) 40.Agricultural cultivator/land holder house wife( )
18. **Education:** 1. Illiterate ( ) 2. Primary School) 3. Secondary school ( ) 4.Higher secondary school 5.Graduate ( )6. Postgraduate ( )
19. **Marital Status:** 1. Married ( ) 2.Unmarrried/Single ( ) 3.Divorse ( ) 4. Widow ( )
20. **Habit of alcohol use:** Habitual ( ) Social ( ) Never ( ) Past history of alcohol consumption ( )
21. **Habit of smoking:** Current smoker ( ) Past smoker ( ) Never ( )
22. **Tobacco use (chewing):** Past ( ) Current use ( ) Never ( )
23. **Body Mass Index (BMI):**
24. **If Tribal: Types of community and language:**
25. **Family type:**

1. Joint ( ) 2. Nuclear ( ) 3. Extended ( )
26. **House type:**

1. Kachha ( ) 2. Semipucca ( ) 3. Pucca ( )
27. **Sanitary facility:** 1. Flush ( ) 2. Kachha ( )3. Open ( )
28. **Nature of use of drinking water:** 1. Filter or boil ( ) 2. Nothing ( )
29. **Education of mother status:** 1. Illiterate ( ) 2. Primary ( ) 3. Above primary **( )**
30. **Mothers employment:** 1. Home maker ( ) 2. Employed **( )**
31. **Father’s Education:** 1. Primary ( ) 2. Above primary ( ) 3. Illiterate **( )**
32. **Fathers occupation:** 1. Govt. ( ) 2. Private ( ) 3. Business ( ) 4. Manual worker ( ) 5. Farmer ( ) 6. Unemployed ( )
33. **Family Size:**
34. **Name of tribes and primitive tribal groups (PVTGs)** : Bagata ( ), Baiga ( ), Banjara ( ), Bathudi ( ), Bhottada( ), Bhuiya ( ), Bhumia ( ), Bhumij ( ), Bhunjia ( ), Binjhal ( ), Binjhia ( ), Birhor ( ), Bondo poraja ( ), Chenchu ( ), Dal ( ), Desia Bhumij ( ), Dharua ( ), Didayi ( ), Gadaba ( ), Gandia ( ), Ghara ( ), Gond ( ), Ho ( ), Holva ( ), Jatapu ( ), Juang ( ), Kandha Gauda ( ), Kawar ( ), Kharia ( ), Kharwar ( ), Khond ( ), Kisan ( ), Kol ( ), Kolah Loharas ( ), Kolha ( ), Koli ( ), Kondadora ( ), Kora ( ), Korua ( ), Kotia ( ), Koya ( ), Kulis ( ), Lodha ( ), Madia ( ), Mahali ( ), Mankidi ( ), Mankirdia ( ), Matya ( ), Mirdhas ( ), Munda ( ), Mundari ( ), Omanatya ( ), Oraon ( ), Parenga ( ), Paroja ( ), Pentia ( ), Rajuar ( ),Santal ( ), Saora ( ), Shabar ( ), Sounti ( ), Tharua ( ), Bondo ( ), Chuktia Bhunjia ( ), Dongria Khond ( ), Kutia Khond ( ), Lanjia Saora ( ), Mankidia ( ), Paudi Bhuyan ( ), Others ( ).
35. **Sunlight exposure:**
36. **Skin color pigmentation:** 1. Dark ( ) 2. Mid Brown ( ) 3. Light ( )

### Dietary and Nutrition Catalogue -2

1. **Food insecurity:** Yes or No

**&Types of food:**

**Cereals**: 1. Rice ( ) 2. Finger Millets ( ) 3.Wheat ( ) 4.Bajra ( ) 5.Jowar ( ) 6. other millets ( )

**Pulses and Legumes:** 1. Red gram ( ) 2. Black gram 3. ( ) Pigeon pea ( ) 4. Green gram ( ) 5. Soyabean 6. ( ) Bangle gram ( ) 7.Soyabean ( ) 8. Pigeon pea ( ) 9.Peas ( ) 10. Any other ( )

**Green leafy vegetables:** 1. Amaranth( ) 2.Spinach 3. ( )Fenugreek leaves ( ) 4. Drumstick leaves ( ) 5.Cabbage ( ) 6.Cauliflower ( ) 7.Coriander leaves ( ) 8. Radish leaves ( ) 9.Curry leaves ( ) 10. Any other ( )

**Vegetables:**1. Bottle gourd ( ) 2. Brinjal ( )3.Cauliflower ( ) 4.Cucumber ( )5.Ladies finger ( ) 6.Tomato ( )7. Cluster bean ( ) 8. Bitter gourd ( ) 9.Double bean ( )10.Ridge gourd( ) 11. Spine gourd ( ) 12. Mushrooms ( ) 13. Any other vegetables ( )

**Roots and Tubers:** 1.Beet root ( ) 2. Carrot ( ) 3. Ginger ( ) 4.Onion ( ) 5.Potato ( ) 6. Radish ( ) 7. Sweet potato ( ) 8.Any other ( )

**Non vegetables:** 1. Flesh ( ) 2.Fish ( )3.Egg ( ) 4.Chicken ( )

**Milk and Oils: 1.** Milk ( ) 2. Curd ( )3. Fruits ( ) 4.Preserved foods ( ) 5.Packed foods ( ) 6.Oils (Palm oil) ( ) 7.Oils (Other oils) ( ) 8.Tea without milk ( ) 9.Tea with milk( ) 10. Ambali ( ) 11. Snacks ( ) 12. Pickles ( ) 13.Peanuts ( ) 14. Lemon Juice ( ) 15.Sugar and Jaggery ( ) 16. Nuts ( )

**Fruits (English/ Hindi):**

**Micronutrients: (Types of Vitamins and Supplements):**

**Macronutrients:**

**Types of salt used:** 1. Iodised ( ) 2. Non-iodised ( )

### Economic Condition Catalogue -3

**Economic status:**

1. Above poverty line [ ] 2. Below poverty line [ ]

**Per capita monthly income (Indian rupees):**

1. 1001-2000 ( ) **2.** 2000-3000 ( ) **3.** 3000-4000 ( ) **4. >**5000 ( ) 5. >10,000( )
2. **Wealth Index:** 1. Poorest ( ) 2.Poor ( ) 3.Middle ( ) 4.Richer ( ) 5.Richest ( )

**Continuous earnings throughout year: Debts condition:**

### Quality on treatment of life Catalogue-4

1. **Sample collected**: Whole blood /Serum/Plasma/ Sputum
2. **History of Signs and Symptoms:**

|  |  |  |  |
| --- | --- | --- | --- |
| a. Fever | Yes/No | b. Diarrhea | Yes/No |
| c. Weightlessness | Yes/No | d. Vomiting | Yes/No |
| e. Expectoration | Yes/No | f. Difficulty in swallowing | Yes/No |
| g. Loss of appetite | Yes/No | h. Itching | Yes/No |
| i. Convulsion | Yes/No | j. Skin rash | Yes/No |
| k. Headache | Yes/No | l. Pulse | Yes/No |
| m. Blood pressure | Yes/No | n. Oral candidiasis | Yes/No |
| o. Generalized weakness | Yes/No | p. Cough | Yes/No |
| q. If yes duration of cough | days |  |  |

**3. Patient follow-up visits:**

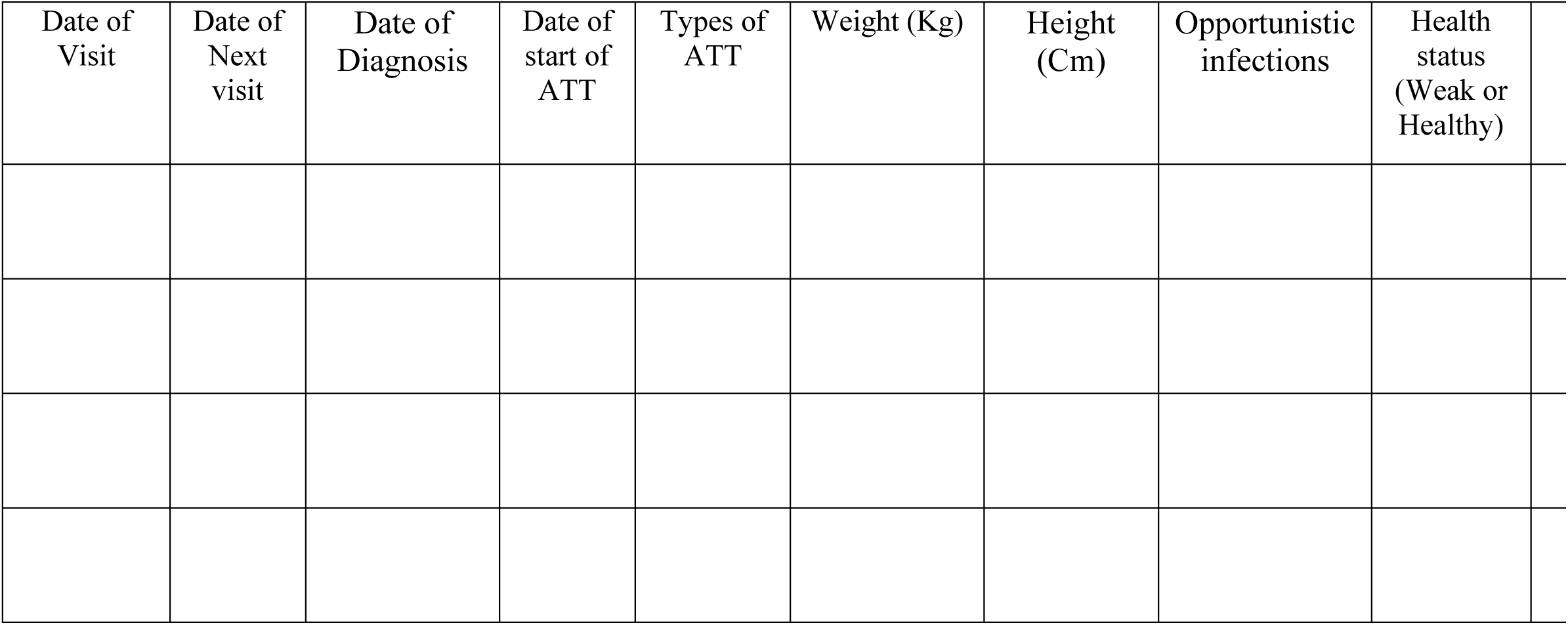
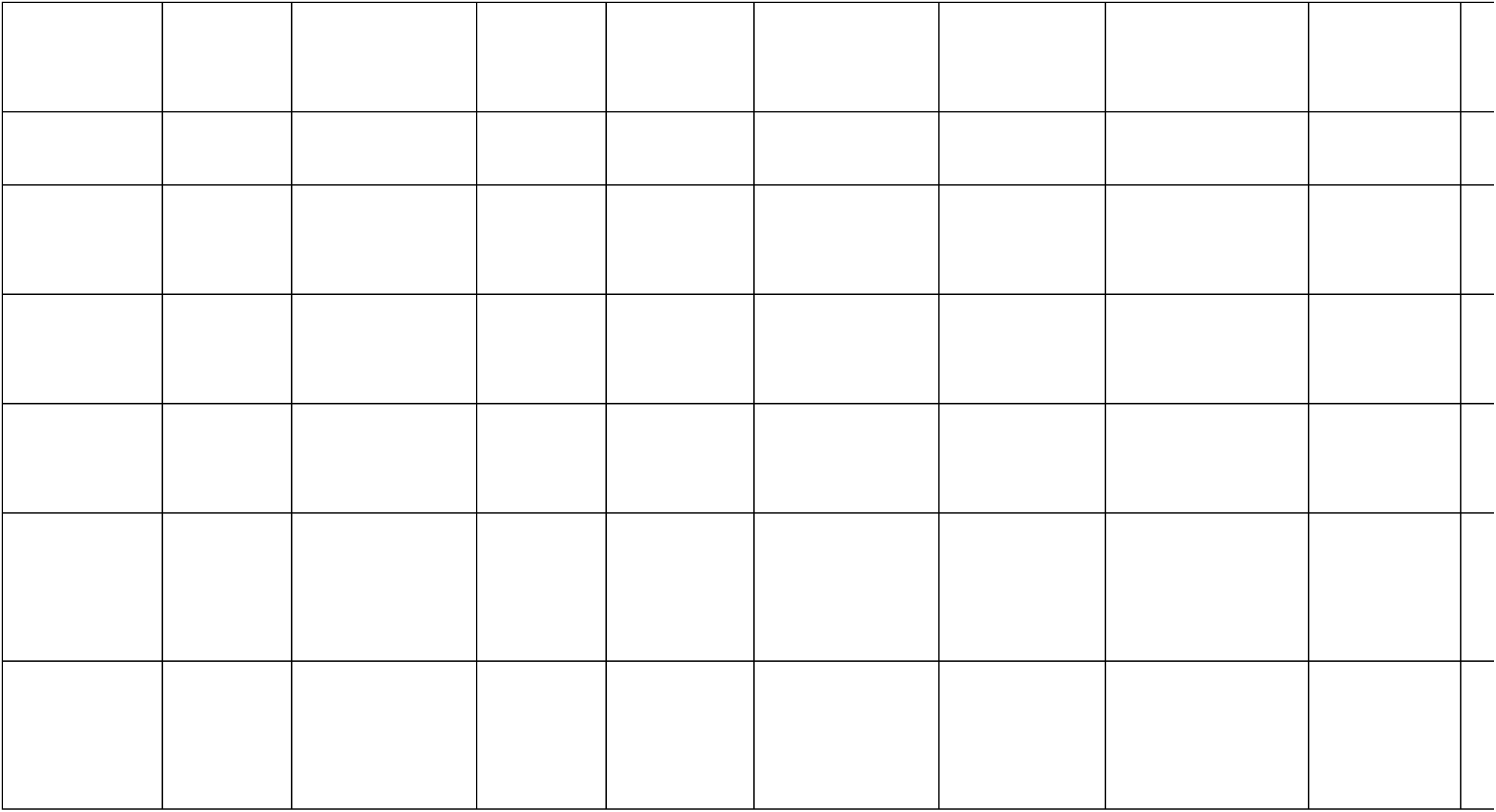

**4. Query on Quality of Life of TB patients**

| **Survey of the Life of Tb patients** | **Response** |  |  |
| --- | --- | --- | --- |
| What is your quality of life after TB? | Good | Bad | Poor |
| Are you satisfied in your present condition of health? | Yes | No |  |
| Do you have now rich energy for working condition? | Yes | No |  |
| Are you satisfied performing your daily activity? | Yes | No |  |
| Are you satisfied your condition of life style? | Yes | No |  |
| Do you have enough money to meet your needs? | Yes | No |  |
| Are you satisfied in your present family condition? | Yes | No |  |
| Are you satisfied in present earning condition? | Yes | No |  |

**5. Query on TB related Stigma**

| **TB Stigma Indicators** | **Desires** |  |
| --- | --- | --- |
| Asked to stay away from work, group | Yes | No |
| Problem with relative to marry | Yes | No |
| Any problem in Marriage | Yes | No |
| Partners refuses sex | Yes | No |
| Any support from spouse | Yes | No |
| Any Problem with children's | Yes | No |
| Desires to keep safe from others | Yes | No |
| Confidant on TB statement | Yes | No |
| Embarrassed or shamed | Yes | No |
| Childrens problem | Yes | No |
| Social burden | High | Low |
| Cultural status of living | High | Low |
| Economic burden | High | Low |
| Daily dietary practice | Yes | No |
| Micro/Macro nutrition supplement | Yes | No |
| Alcohol/Fruit beverages | Yes | No |
| Vitamin D uptake | Yes | No |
| Quality of food | Good | Bad |
| Millet's food taken | Yes | No |

Patient signature:
Student signature:
Date of collection:

