## Supplementary material for "Socio-cultural practices, dietary and nutrition patterns, economic and vitamin-D deficiency status of pulmonary tuberculosis patients (PTB) of tribal and urban population of India: An Explanatory Model Interview Catalogue (EMIC)": Ethics Certificate (from another version of this preprint: version 1, 2023-07-26)

आई.सी.एम.आर - क्षेत्रीय आयुर्विज्ञान अनुसंधान केन्द्र

icmr INDIAN COUNCIL OF MEDICAL RESEARCH | RMRCBB REGIONAL MEDICAL RESEARCH CENTRE, BHUBANESWAR

### ICMR - REGIONAL MEDICAL RESEARCH CENTRE, BHUBANESWAR

#### INSTITUTIONAL HUMAN ETHICS COMMITTEE

IRB No: ECR/911/Inst/OR/2017
Federalwide Assurance # FWA00027286

***Protection of Human Subjects***
***IEC Certification/ Declaration of Exemption***

Chairman
**Prof. Dr. Jadunath Prasad Das**
Clinician

Member
**Prof. Aruna Kumari Misra**
Basic Scientist, Life Science
**Prof. Dr. Prasanna Kumar Dash**
Medical Scientist
**Mrs. Kasturika Patnaik**
Socially Aware Person
**Dr. Pramod Kumar Acharya**
Clinician
**Dr Sisir Kumar Mahapatra**
Clinician
**Sri Santanu Das**
Legal Expert
**Prof. Rita Ray**
Social Scientist
**Prof. Dr. Sudhanshu Sekhar Mishra**
Basic Medical Scientist
**Dr. Anna S. Kerketta**
Basic Medical Scientist

Ref: …*ICMR-RMRC/IHEC-2022/118* Date: 25/07/2022

**1.** Request Type
- ☐ √ Original
- ☐ Continuation
- ☐ Exemption

**2.** Type of Programme
- ☐ √ Research
- ☐ Contract Research
- ☐ Collaborative
- ☐ Fellowship
- ☐ Others
- ☐ Institutional Activity-Out Patient Care/ Surveillance/ Referral & Outbreak Investigation

**2.** Proposal Identification No.

- Institutional ID:
- Funding Agency & ID (If any):

**3.** Title of Research or Activity: *A comparative study of the Vitamin D-Deficiency, Vitamin-D receptor polymorphisms in Pulmonary Tuberculosis Patients in tribes and urban population of Odisha, India*

**4.** Principal Investigator: *Dr Jyotirmayee Turuk, Scientist C*

**5.** Key Personnel at RMRC Bhubaneswar & Role:

**6.** Certification of IRB review: This activity/ proposal has been reviewed and approved by the IEC in accordance with the Indian Council for Medical Research (ICMR), guidelines and other GCP recommendations, by full IRB review or expedited review in July, 2022.

**7.** Remarks (If any):

- Confidentially to be maintained
- Consent to be obtained

**8.** This activity/ proposal has been scrutinized and declared for exemption for review by IEC as there is less than minimal risk and no linked identifiers.

Dr. Sidhartha Giri
Scientist -E (Medical)
Indian Council of Medical Research-
Regional Medical Research Center
Bhubaneswar-751023

Sidhartha Giri

**Dr. Sidhartha Giri**
**Scientist- E**
**Member Secretary** Signature:

Name & Designation of the Official Date: 25/07/2022
